# Causal associations between gut microbiota and cancers

**DOI:** 10.1101/2025.02.17.25322359

**Authors:** Zihan Yue, Junwei Yan, Linyuan Shen, Chunguang Zhao, Xiaopeng Yang, Yiying Yao, Dongmei Song, Chenyang Xu, Chenchen Bi, Zhongkui Xiong, Hongli Ma, Zheng Liu

**Affiliations:** Department of Nursing, School of Medicine, Shaoxing University, Shaoxing, Zhejiang, China; Department of Pharmacology, School of Medicine, Shaoxing University, Shaoxing, Zhejiang, China; Department of Blood Transfusion, Affiliated Hospital of Shaoxing University, Shaoxing, Zhejiang, China; Zhejiang Chint Electrics Co.,Ltd, Wenzhou, Zhejiang, China; Department of Radiotherapy, Shaoxing Sencond Hospital, Shaoxing, Zhejiang, China; Department of Nursing, Shaoxing People’s Hospital, Shaoxing, Zhejiang, China

**Keywords:** Gut microbiota, cancers, Mendelian randomization, meta-analysis, causal relationship

## Abstract

**Background:** Emerging evidence suggests that the gut microbiota is associated with various cancer-related outcomes. Recent studies using Mendelian randomization (MR) have indicated a causal relationship between gut microbiota and several types of cancer. However, these conclusions remain controversial. To clarify this relationship, we conducted a systematic review and meta-analysis of MR studies to investigate the potential causal links between gut microbiota and cancer.

**Method:** In this study, we searched PubMed, Embase, the Cochrane Library, and Web of Science up to May 2024 for eligible MR studies and performed meta-analyses.

**Results:** Our findings indicate that genetically determined gut microbiota is associated with 12 different types of neoplastic outcomes. We identified nine genera of gut microbiota that are generally associated with a reduced risk of developing cancer. However, we also found that the order Mollicutes RF9 (OR = 1.037, 95% CI 1.013-1.060) may be a potential risk factor for breast and lung cancer. Furthermore, we discovered that the families Alcaligenaceae (OR = 1.034, 95% CI 1.000-1.068), Enterobacteriaceae (OR = 1.077, 95% CI 1.020-1.135), and Lactobacillaceae (OR = 1.033, 95% CI 1.008-1.059) are negatively associated with the risk of breast cancer. On the other hand, the genetically predicted family Lactobacillaceae (OR = 0.892, 95% CI 0.848-0.937), along with other families and genera such as Veillonellaceae, Coprococcus, Dorea, Eubacterium, Lachnospiraceae, Ruminococcus, Ruminococcaceae, and FamilyXIII, were found to be correlated with a lower risk of lung cancer. Additionally, the family Peptostreptococcaceae, Veillonellaceae, Streptococcaceae, and certain genera including Eubacterium, Lachnospiraceae, and Ruminococcaceae, were identified as protective factors against glioblastoma. The evidence from published MR studies supports the notion that the gut microbiota plays a causal role in various neoplastic diseases.

**Conclusions:** In conclusion, there is a causal relationship between gut microbes and cancer, and it is related to microbial species and tumor type. Further research is necessary to understand the underlying mechanisms and to explore the potential for using gut microbiota in prediction, prevention, and therapeutic strategies for these diseases. Studying changes in the microbiome in cancer has significant implications for developing noninvasive diagnostic tools and innovative interventions that could alter the progression of these diseases.

## Introduction

The human gut is home to approximately 3.8 trillion microorganisms, collectively weighing around 1.8 kilograms, and are collectively known as the gut microbiota. These microorganisms play a crucial role in maintaining health and physiological balance, influencing metabolism and modulating the immune system^1^. Recent research has highlighted the significant impact of the gut microbiota on cancer development, with different types of tumors potentially harboring unique microbial communities^2^. The gut microbiota within the tumor microenvironment is believed to actively contribute to cancer development and progression through various mechanisms, including promoting cell proliferation, evading growth-inhibiting mechanisms, resisting apoptosis, facilitating angiogenesis, and subverting immune surveillance^3^.

Advancements in preclinical research have revealed the complex pathways through which microbes can influence the effectiveness of cancer treatments. This has been further supported by clinical trials, which have underscored the therapeutic potential of modulating the microbiota in oncology^4^. The intricate relationship between the gut microbiota and various cancer therapies—ranging from chemotherapy^5^, and radiotherapy^6^, to targeted therapy^7^ and immunotherapy^8^ is now a well-documented factor in the field.

Unlike the fixed nature of host genetics, the adaptability of the microbiota offers a unique advantage. It can be modified through a range of strategies, including fecal microbiota transplantation (FMT), the use of probiotics, and targeted antibiotic therapies. This adaptability is paving the way for a more personalized approach to cancer treatment, marking a significant shift towards tailored therapies in the realm of personalized medicine^4^. Most previous research conclusions regarding the relationship between gut microbiota and cancers have been based on observations of the microbiota’s composition and changes in patients’ fecal samples, as well as the outcomes of trials involving the transplantation of gut microbiota into gnotobiotic mice^9–12^. However, the causal relationship between cancer and microbiota remains to be fully elucidated.

MR is an instrumental variable analysis that mitigates bias from reverse causality^13^ and addresses major limitations inherent in observational studies, such as unmeasured confounding, ascertainment bias, and small sample sizes^14,15^. It serves to test the causal relationship between the microbiota and cancer risk^16^. However, several MR studies have presented contradictory conclusions, particularly when characterizing the association between microorganisms and tumors. Therefore, to thoroughly evaluate and synthesize the evidence concerning the causal role of gut microbiota in various cancers, we undertook a systematic review and meta-analysis of published MR studies. Investigating the specific microbiota associated with different cancers offers novel insights and strategies for cancer prevention, treatment, and health management.

## Methods

### Search strategy and selection criteria

We conducted an exhaustive literature search in the Cochrane, PubMed, Embase, and Web of Science databases from inception through May 31, 2024, to identify all MR studies exploring the association between genetically determined gut microbiota and cancer outcomes. The search terms included: “Gastrointestinal Microbiomes,” “Gut Microbiome,” “Intestinal Microbiota,” or “Microbiome” for the exposure; “Tumor,” “Neoplasm,” or “Cancer” for the outcome; and “Mendelian randomization” for the study design. The detailed search strategy is presented in the Supplementary Method. Additionally, we identified potentially eligible studies through the reference lists of included studies and relevant reviews.

Inclusion criteria for studies were: (1) utilization of MR; (2) use of gut microbiota as the exposure variable; and (3) measurement of the causal association between the exposure and one or more cancer outcomes. Studies were excluded if they met any of the following criteria: (1) non-human studies; (2) conference abstracts, editorials, or reviews; (3) those not written in English or Chinese; or (4) if the full text was not available. In cases where multiple studies reported the same outcome in the same population, we prioritized the study with the largest participant sample.

After the literature search, all articles were imported into NoteExpress, and duplicate records were eliminated. Two independent reviewers assessed the articles based on the predefined inclusion and exclusion criteria, with any disagreements resolved by a third reviewer. The selection process began with titles and abstracts, followed by a full-text review.

### Data extraction and quality assessment

Data extracted from each study included the surname of the first author and the publication year. We also recorded the consortium or study that provided the genetic variants for gut microbiota exposure and the one that supplied the genetic association estimates for cancers. Additionally, we noted the sample size, defined as the number of cases and non-cases. We extracted the relative risk estimate (reported as the odds ratio [OR]) along with the corresponding 95% confidence interval (CI) for the association of gut microbiota. One investigator was responsible for extracting the data, and four other investigator verified the accuracy of these data. Two researchers assessed the quality of the research by adopting a modified version of the Strengthening the Reporting of Observational Studies in Epidemiology using Mendelian Randomization (STROBE-MR) guidelines. The guidelines were adapted based on articles reporting quality assessment approaches used to document MR studies. The results from the evaluation of study quality are provided in Additional file 1: Table S1.

### Statistical analysis

Stata (version 17.0, Stata Corporation) software was utilized to conduct the meta-analysis of the included MR associations. The odds ratios (ORs) per 1 standard deviation (SD) increase in gut microbiota, with their respective 95% CIs, were pooled for each cancer outcome using a random-effects model.

## Results

### Literature search and study selection

The literature search yielded 223 records, of which 71 original articles reported findings on genetic liability to gut microbiota in relation to one or more specific outcomes. After excluding studies that used overlapping or identical outcome data, 58 articles based on non-overlapping populations were deemed eligible for inclusion in one or more meta-analyses^17–30^. Figure 1 provides a summary of the study selection process and the designs of the included studies.

**Figure 1.**
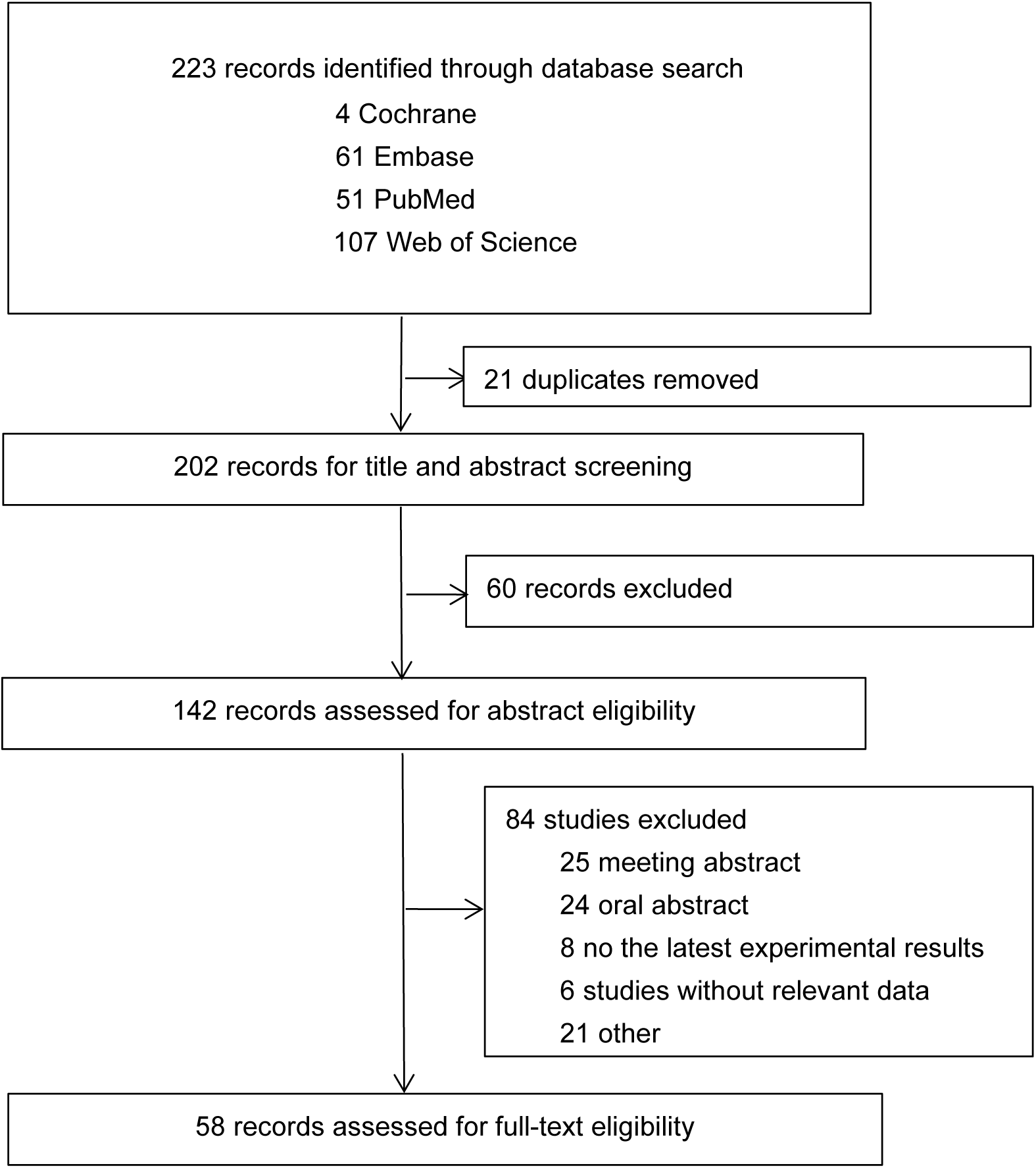
Flow chart of the systematic review and meta-analysis.

### Causal relationship between cancers and gut microbiota at phylum, class and order level

At the phylum level, we identified nine associations with cancers. Through meta-analysis, we combined these findings and determined that the phyla Proteobacteria (OR = 0.963, 95% CI 0.935-0.990), Tenericutes (OR = 0.968, 95% CI 0.941-0.996), and Verrucomicrobia (OR = 0.983, 95% CI 0.969-0.997) were associated with a reduced risk of cancer. However, we found no significant associations between the other phyla and cancer. Under the phylum Proteobacteria, classes Alphaproteobacteria (OR = 0.977, 95% CI 0.964-0.990), Gammaproteobacteria (OR = 0.967, 95% CI 0.939-0.995) and the order Rhodospirillales (OR = 0.963, 95% CI 0.937-0.989) were associated with a decreased risk of cancers. Under the phylum Tenericutes, the class Mollicutes (OR = 0.967, 95% CI 0.938-0.995) was associated with a decreased risk of cancers. However, the order MollicutesRF9 (OR = 1.037, 95% CI 1.013-1.060) was associated with an increased risk of cancers. Under the phylum Verrucomicrobia, the class Verrucomicrobiae (OR = 0.983, 95% CI 0.972-0.994) and the order Verrucomicrobiales (OR = 0.988, 95% CI 0.980-0.997) were associated with a decreased risk of cancers. (See Figure 2)

**Figure 2.**
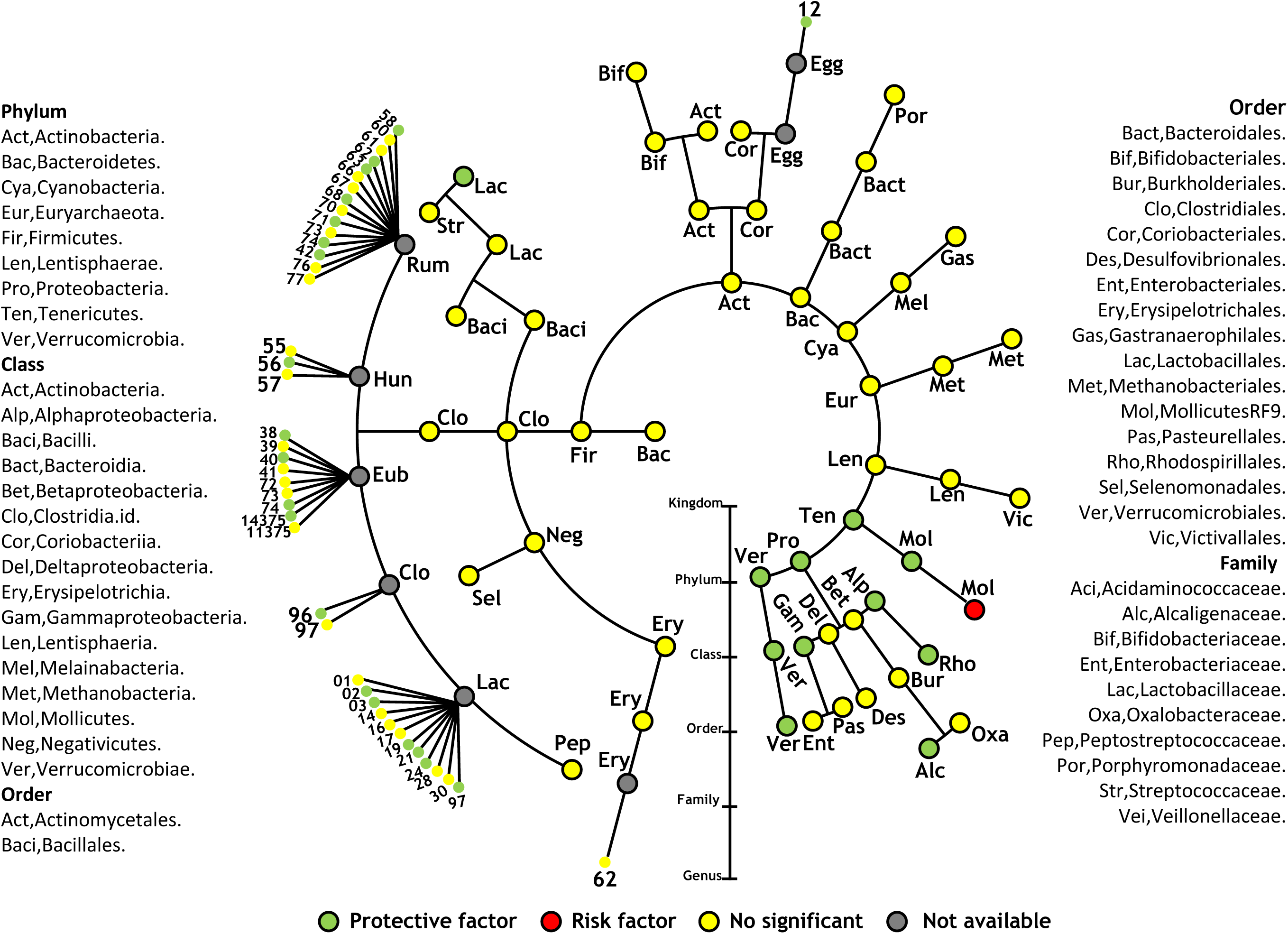
Cladogram of odds ratio (OR) of the association between gut microbiome and the risk of cancer outcomes. From the inner ring to the outer ring are the kingdom, phylum, class, order, family and genus.

Since we have explored some meaningful results in phyla, classes, and orders of taxa respectively, we continue to explore the levels of intestinal flora at the family and genus levels. However, due to the large amount of data, we analyzed the top ten families and genera with the highest data volume, respectively. Interestingly, most of the families and genera are concentrated in the order Clostridiales under the Firmicutes taxa.

Under the order Clostridiales, the family *Ruminococcaceae* (OR = 0.997, 95% CI 0.995-0.998) was associated with a decreased risk of cancers. Among them, *Ruminococcaceae* is derived from the aggregate effect of its subtaxa. Under other orders, the families *Alcaligenaceae.id.2875* (OR = 0.963, 95% CI 0.937-0.989) and *Lactobacillaceae.id.1836* (OR = 0.948, 95% CI 0.921-0.975) were associated with a decreased risk of cancers. Under the family *Eubacterium* taxa, the genera *Eubacterium brachy group.id.11296*, *Eubacterium hallii group.id.11338*, *Eubacterium rectale group.id.14374*, *Eubacterium ruminantium group.id.11340*, and *Eubacterium xylanophilum group.id.14375* were associated with a decreased risk of cancers. Under the family *Hungateiclostridiaceae* taxa, the genus *Ruminiclostridium 6.id.11356* was associated with a decreased risk of cancers, while the genus *Ruminiclostridium* 9*.id.11357* was associated with an increased risk of cancers. Under the family *Lachnospiraceae* taxa, several genera including *Coprococcus 2.id.11302*, *Coprococcus 3.id.11303*, *Dorea.id.1997*, *Lachnospiraceae FCS020 group.id.11314*, *Lachnospiraceae NK4A136 group.id.11319*, *Lachnospiraceae UCG001.id.11321*, and *Lachnospiraceae UCG004.id.11324* were associated with a decreased risk of cancers. The genus *Lachnospiraceae UCG010.id.11330* was associated with an increased risk of cancers. Under the family *Ruminococcaceae* taxa, several genera including *Ruminococcaceae NK4A214 group.id.11358*, *Ruminococcaceae UCG004.id.11362*, *Ruminococcaceae UCG005.id.11363*, *Ruminococcaceae UCG011.id.11368*, *Ruminococcaceae UCG014.id.11371*, *Ruminococcus 2.id.11374*, and *Ruminococcus gauvreauii group.id.11342* were associated with a decreased risk of cancers. The genus *Ruminococcaceae UCG010.id.11367* was associated with an increased risk of cancers. Under other family taxa, the *Family XIII AD3011 group*.id.11293 and *Adlercreutzia.id.812* were associated with a decreased risk of cancers. Because this is a causal study that excludes confounding factors, even though the taxa are related on the branch plot, they are all statistically independent. (See Figure 2)

### Causal effects of gut microbiota in the family and genera level on the ten cancer types

At the phylum level, high heterogeneity between studies was only observed in the analyses of the phyla Actinobacteria and Verrucomicrobia. We analyzed them in subgroups by cancer type and found that the heterogeneity of Actinobacteria was mainly concentrated in thyroid cancer and digestive cancers, significantly reducing other heterogeneity. It was found to be a protective factor against digestive cancers (OR = 0.81, 95% CI 0.625, 0.996), thyroid cancer (OR = 0.634, 95% CI 0.470, 0.799), gynecological cancers (OR = 0.998, 95% CI 0.997, 0.999), and lymphoma (OR = 0.819, 95% CI 0.656, 0.981). This result may indicate insufficient data volume, necessitating further analysis. In Verrucomicrobia, heterogeneity was significantly reduced, and it was found to be a protective factor against lymphoma (OR = 0.765, 95% CI 0.632, 0.897) and lung cancers (OR = 0.862, 95% CI 0.796, 0.928), but a risk factor for glioblastoma (OR = 1.268, 95% CI 1.07, 1.466). (Table 2) To further analyze the causal relationship between gut microbiota and specific cancers, we selected the top ten families and genera based on data volume for further analysis and found that different cancers had closer relationships with specific families and genera. (Figure 3)

**Figure 3.**
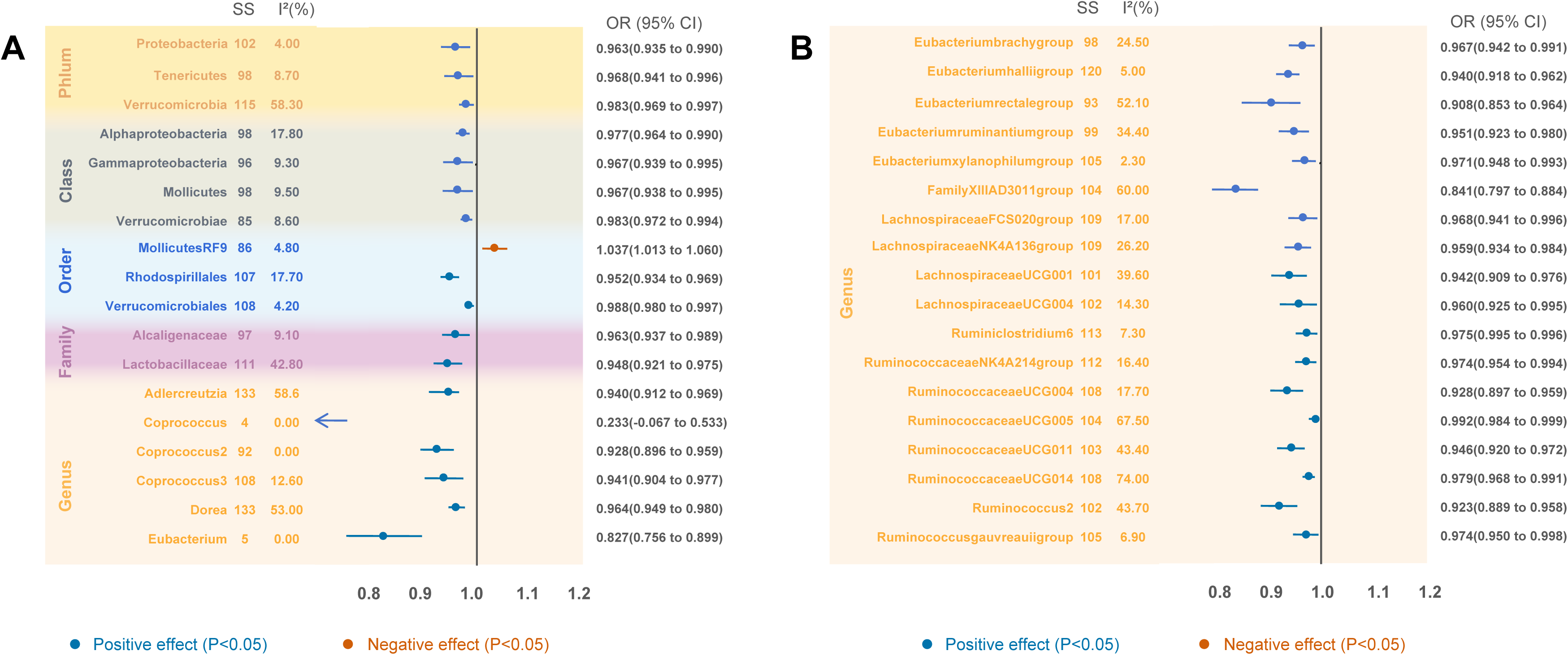
Forest plot of meta-analysis results of the association between gut microbiome and risk of cancer. The effect on the x-axis is the odds ratio of cancer per 1 standard deviation change in the exposure.

**Figure 4.**
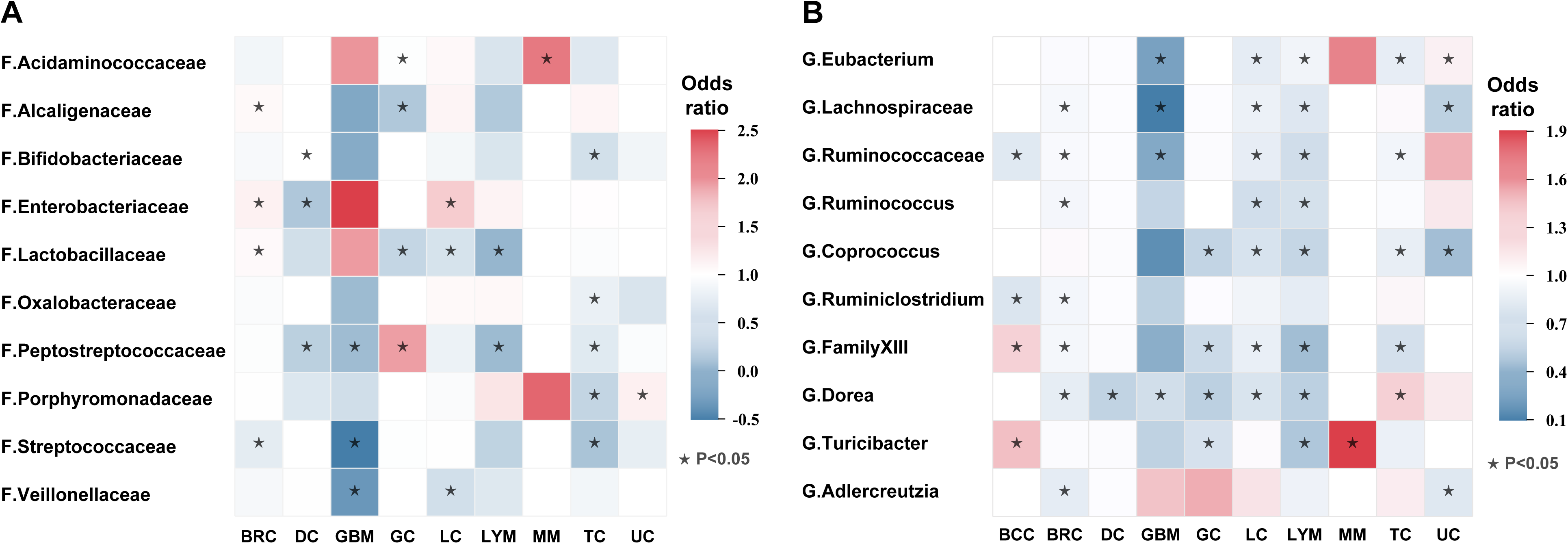
Heatmap of the ORs. Color in the heatmap indicates the OR of gut metabolite and cancers. Darker red color indicates higher OR values. The asterisk indicates statistically significant OR. BCC, Basal Cell Carcinoma; BRC, Breast Cancer; DC, Digestive Cancers; GBM, Glioblastoma; GC, Gynecological cancer; LC, Lung Cancer; LYM, Lymphoma; MM, Multiple Myeloma; TC, Thyroid Cancer; UC, Urinary Cancers

#### Basal Cell Carcinoma

The genus *Ruminococcaceae* (OR = 0.887, 95% CI 0.786-0.988) and genus *Ruminiclostridium* (OR = 0.864, 95% CI 0.744-0.984) were associated with a decreased risk of basal cell carcinoma. Conversely, the genus *Family XIII* (OR = 1.124, 95% CI 1.001-1.247) and genus *Turicibacter* (OR = 1.178, 95% CI 1.064-1.291) were associated with an increased risk of basal cell carcinoma.

#### Breast Cancer

The family *Streptococcaceae.id.1850* (OR = 0.929, 95% CI 0.899-0.959), the genus *Adlercreutzia* (OR = 0.907, 95% CI 0.866-0.949), genus *Dorea* (OR = 0.911, 95% CI 0.871-0.951), genus *FamilyXIII* (OR = 0.970, 95% CI 0.945-0.995), genus *Lachnospiraceae* (OR = 0.976, 95% CI 0.966-0.987), genus *Ruminococcus* (OR = 0.968, 95% CI 0.950-0.986), genus *Ruminococcaceae* (OR = 0.987, 95% CI 0.982-0.991) and genus *Ruminiclostridium* (OR = 0.965, 95% CI 0.947-0.983) were associated with a decreased risk of breast cancer. Conversely, the family *Alcaligenaceae.id.2875* (OR = 1.034, 95% CI 1.000-1.068), the family *Enterobacteriaceae.id.3469* (OR = 1.077, 95% CI 1.020-1.135) and family *Lactobacillaceae.id.1836* (OR = 1.033, 95% CI 1.008-1.059) were associated with an increased risk of breast cancer.

#### Digestive cancers

The family *Enterobacteriaceae.id.3469* (OR = 0.786, 95% CI 0.637-0.935), family *Peptostreptococcaceae.id.2042* (OR = 0.818, 95% CI 0.711-0.925), and genus *Dorea* (OR = 0.756, 95% CI 0.582-0.929) were associated with a decreased risk of digestive cancers. The family *Bifidobacteriaceae.id.433* (OR = 1.001, 95% CI 1.000-1.002) was associated with an increased risk of digestive cancers.

#### Glioblastoma

The family *Peptostreptococcaceae.id.2042* (OR = 0.742, 95% CI 0.638-0.846), family *Veillonellaceae.id.2172* (OR = 0.389, 95% CI −0.187-0.965), family *Streptococcaceae.id.1850* (OR = 0.231, 95% CI −0.230-0.692), the genus *Eubacterium* (OR = 0.455, 95% CI 0.216-0.693), genus *Lachnospiraceae* (OR = 0.238, 95% CI 0.018-0.459) and genus *Ruminococcaceae* (OR = 0.500, 95% CI 0.255-0.745) were associated with a decreased risk of glioblastoma.

#### Gynecological cancers

The genus *Coprococcus* (OR = 0.761, 95% CI 0.655-0.867), genus *Dorea* (OR = 0.735, 95% CI 0.667-0.804), genus *Family XIII* (OR = 0.799, 95% CI 0.745-0.854) and genus *Turicibacter* (OR = 0.842, 95% CI 0.789-0.896) were associated with a decreased risk of gynecological cancers.

#### Lung Cancer

The family *Lactobacillaceae.id.1836* (OR = 0.892, 95% CI 0.848-0.937), family *Veillonellaceae.id.2172* (OR = 0.878, 95% CI 0.830-0.926), the genus *Coprococcus* (OR = 0.850, 95% CI 0.800-0.900), genus *Dorea* (OR = 0.856, 95% CI 0.761-0.951), genus *Eubacterium* (OR = 0.908, 95% CI 0.884-0.933), genus *Lachnospiraceae* (OR = 0.937, 95% CI 0.909-0.964), genus *Ruminococcus* (OR = 0.824, 95% CI 0.783-0.866), genus *Ruminococcaceae* (OR = 0.913, 95% CI 0.890-0.937) and genus *Family XIII* (OR = 0.923, 95% CI 0.856-0.990) were causally associated with lung cancer. However, the OR values were all less than 1, suggesting a potential tumor suppressor effect. Conversely, the family *Enterobacteriaceae.id.3469* (OR = 1.267, 95% CI 1.112-1.423) was associated with an increased risk of lung cancer.

#### Lymphoma

The genus *Coprococcus* (OR = 0.775, 95% CI 0.669-0.881), the genus *Dorea* (OR = 0.737, 95% CI 0.480-0.994), genus *Eubacterium* (OR = 0.955, 95% CI 0.915-0.996), genus *Family XIII* (OR = 0.626, 95% CI 0.522-0.729), genus *Lachnospiraceae* (OR = 0.878, 95% CI 0.826-0.930), genus *Ruminococcus* (OR = 0.840, 95% CI 0.776-0.903), genus *Ruminococcaceae* (OR = 0.818, 95% CI 0.771-0.864) and genus *Turicibacter* (OR = 0.683, 95% CI 0.585-0.781) were associated with a decreased risk of lymphoma.

#### Oropharyngeal Cancer

The genus *Dorea* (OR = 1.004, 95% CI 1.001-1.007) and genus *Ruminococcaceae* (OR = 1.004, 95% CI 1.001-1.007) were associated with an increased risk of oropharyngeal cancer.

#### Thyroid Carcinoma

The family *Bifidobacteriaceae.id.433* (OR = 0.881, 95% CI 0.806-0.956), family *Oxalobacteraceae.id.2966* (OR = 0.945, 95% CI 0.893-0.997), family *Peptostreptococcaceae.id.2042* (OR = 0.919, 95% CI 0.846-0.992), family *Streptococcaceae.id.1850* (OR = 0.772, 95% CI 0.699-0.845), family *Porphyromonadaceae.id.943* (OR = 0.839, 95% CI 0.715-0.964), the genus *Coprococcus* (OR = 0.918, 95% CI 0.860-0.977), genus *Eubacterium* (OR = 0.913, 95% CI 0.891-0.935) and genus *Family XIII* (OR = 0.836, 95% CI 0.771-0.900) were associated with a decreased risk of thyroid carcinoma. The genus *Dorea* (OR = 1.127, 95% CI 1.011-1.243) was associated with an increased risk of thyroid carcinoma.

#### Urinary cancers

The genus *Adlercreutzia* (OR = 0.883, 95% CI 0.857-0.910), the genus *Coprococcus* (OR = 0.630, 95% CI 0.405-0.855) and genus *Lachnospiraceae* (OR = 0.740, 95% CI 0.560-0.920) were associated with a decreased risk of urinary cancers. The family *Porphyromonadaceae.id.943* (OR = 1.082, 95% CI 1.017-1.146) and genus *Eubacterium* (OR = 1.037, 95% CI 1.001-1.073) were associated with an increased risk of urinary cancers.

## Discussion

In this systematic review and meta-analysis of MR studies, we have conducted the most extensive analysis to date, encompassing 58 studies and 22,818 data. Utilizing taxonomical methods, we have delved into the causal relationship between gut microbiota and the spectrum of cancers. Our findings reveal that the phyla Proteobacteria, Tenericutes, and Verrucomicrobia exert protective effects against cancer, while the order MollicutesRF9.id.11579 emerges as a potential risk factor for breast and lung cancer. Furthermore, we identified the family *Alcaligenaceae, Lactobacillaceae*, and genera *Adlercreutzia, Dorea, Eubacterium* brachy group*, Lachnospiraceae* FCS020 group, and *Ruminococcaceae* UCG004 as being associated with a reduced cancer risk. Expanding our investigation to the phylum level, Actinobacteria demonstrated a protective role in digestive, thyroid, gynecological cancers, and lymphoma. Intriguingly, Verrucomicrobia showed a protective effect for lung cancer and lymphoma, yet not for glioblastoma, underscoring the nuanced influence of microbial taxa on cancer risk.

At the familial and generic levels, our analysis uncovered a more complex picture. The family *Peptostreptococcaceae* was identified as a risk factor for gynecological cancers but offered protection against digestive cancers, glioblastoma, and lymphoma. Similarly, the genus *Turicibacter* presented as a risk for basal cell carcinoma and multiple myeloma, yet it was protective for gynecological cancers and lymphoma. Notably, the family *Enterobacteriaceae*, while a risk factor for breast and lung cancers, paradoxically acts as a protective factor against digestive cancers. Moreover, the families *Lactobacillaceae, Veillonellaceae*, and the genus *Eubacterium*, along with *Lachnospiraceae, Ruminococcaceae, Ruminococcus, Coprococcus, Dorea*, and *Turicibacter*, were found to be protective against lung cancer. In contrast, *Enterobacteriaceae* was identified as a risk factor for this disease.

In our comprehensive analysis, the phylum Proteobacteria has emerged as a significant factor in cancer prognosis. Our findings indicate that higher levels of Proteobacteria in pancreatic tumors are associated with extended survival times, positing these bacteria as potentially beneficial in pancreatic cancer contexts^31^, This correlation is mirrored in ovarian cancer, where an increased presence of Proteobacteria is also noted^32^. In the context of pancreatic ductal adenocarcinoma, Proteobacteria may contribute to immune evasion by upregulating immunosuppressive cytokines such as IL-10, which can diminish type 1 helper T cell activity and drive tumor-associated macrophages toward an M2 phenotype, thereby fostering a more immunosuppressive tumor microenvironment^33^. Although preliminary experimental evidence points to a role for gut microbiota in tumor promotion and development^34^, the specific mechanisms through which this occurs have yet to be fully established. Further research is essential to uncover the detailed mechanisms by which Proteobacteria could influence host immune responses and cancer treatment efficacy. The potential mechanisms we propose for investigation include: (1) Microbial Ecosystem Effects: The influence of Proteobacteria on the gut microbiota’s composition and function^35^, (2) Intestinal Barrier Modulation: Impacts on the intestinal epithelium and associated lymphoid tissues, potentially via autophagy and apoptosis induction^36,37^, (3)Pattern-Recognition Receptor Activation: The stimulation of immune receptors by microbial components^38^, (4) Neuroendocrine System Interactions: The influence on neuroendocrine pathways through gut hormone secretion^39^, (5) Systemic Metabolic Impact: The role in systemic metabolism via the synthesis of polyamines and B vitamins^40^, and (6) Cross-Reactive Immune Responses: The potential to induce immune reactions against microbial antigens that mimic tumor-associated antigens^41,42^. These insights underscore the importance of exploring the microbiota’s mechanistic links to cancer. Such knowledge is vital for the development of microbiota-targeted therapeutic strategies that could enhance the efficacy of cancer treatments by leveraging the protective effects of Proteobacteria.

Our MR analysis has identified the phylum Actinobacteria as a protective factor against a range of cancers, including those of the digestive system, thyroid, gynecological organs, and lymphoma. This is further supported by previous findings highlighting a high prevalence of Actinobacteria in breast cancer tissue samples^43^. Notably, the family Bifidobacteriaceae appears to offer protection against digestive and thyroid cancers. Variations in Bifidobacterium levels have been linked to different clinical stages of breast cancer, suggesting a role for the microbiome in disease progression^44^. Consistent with this, elevated Bifidobacterium levels have been observed in patients with colorectal adenomas and in mouse models of advanced pancreatic cancer ^33,45^. Moreover, Bifidobacterium has been correlated with improved responses to anti-PD-L1 therapies^46^. However, it is important to note that while generally beneficial, certain Bifidobacterium species may act as potential pathogens, indicating a complex relationship with human health^47,48^. Ruminococcus, an early-identified gut bacterium with key metabolic functions, including the breakdown of cellulose, has been identified as a protective factor in breast, lung, and lymphoma. Beyond its metabolic role, Ruminococcus contributes to intestinal barrier stabilization, diarrhea prevention, reduction in kidney stone formation, and decreased risk of colorectal cancer^49^. Conversely, the family Peptostreptococcaceae presents a risk for gynecological cancers but interestingly, acts protectively against digestive cancers, glioblastoma, and lymphoma^17^.These findings underscore the intricate and sometimes paradoxical nature of microbial associations with cancer, emphasizing the need for a nuanced understanding of the microbiome’s role in cancer development and progression.

In summary, the gut microbiota’s diverse species exert variable influences on the tumor microenvironment, with potential to be either beneficial or detrimental to health^50^. Our review indicates that the compositional differences in gut microbiota between healthy individuals and those with cancer may not solely indicate causation, but could also be a bystander effect or even contribute to carcinogenesis^51^. Diet significantly shapes the gut microbiota, thereby indirectly influencing cancer risk^52^. Strategically, probiotics offer a practical avenue for modulating the gut microbiota^53^. Prebiotics, which are non-living substances, and probiotics, which are live bacteria, can enhance the growth and activity of beneficial species. For instance, probiotic supplementation with Bifidobacterium lactis and Lactobacillus acidophilus in colorectal cancer (CRC) patients has been shown to increase butyrate production and reduce CRC-promoting bacteria^54^.Such interventions can regulate inflammation, bolster immune responses, and enhance antitumor immunity^55^. Furthermore, the combination of immunotherapy with probiotics represents a promising research frontier, allowing for the development of targeted treatments that leverage the synergistic effects of multiple microorganisms^2^.

This study’s strength lies in its systematic review and meta-analysis of MR studies, which provides a robust investigation into the causal links between gut microbiota and cancer. However, it is not without limitations. The analysis was conducted at the genus level, which does not capture species or strain-level nuances. Additionally, the heterogeneity among the studies included could introduce some bias into our estimates of causality. Lastly, future research should explore the relationship between specific gut microbiota and various cancer subtypes to build upon these findings and advance cancer prevention and treatment strategies.

## Conclusion

In conclusion, this study delivers a sweeping evaluation of the potential causal links between the gut microbiota and the pan-cancer risk landscape. Our findings not only shed new light on the etiology of cancer but also set the stage for leveraging the gut microbiome as a biomarker, heralding a new era in personalized cancer therapy and patient prognosis. By elucidating how the gut microbiota influences tumor onset and progression, our research provides substantial evidence that could facilitate the incorporation of microbiome assessments into cancer management strategies. Early detection through fecal testing could identify high-risk individuals, while microbiota modulation presents a promising preventive and therapeutic avenue. Looking ahead, the integration of gut microbiota analysis into routine cancer surveillance, diagnostics, and therapeutics is poised to transform cancer care, enhancing both its efficacy and personalization.

## Supporting information

Quality assessment

## Data Availability

All data produced in the present study are available upon reasonable request to the authors
All data produced in the present work are contained in the manuscript
All data produced are available online at

## Author Contributions

All authors were involved in the conception of this review, Zihan Yue: collected data, interpreted the results, wrote the manuscript and drew the figures. Zheng Liu: designed and supervised the study and revised the manuscript. Hongli Ma: checked and reviewed the manuscript. Junwei Yan: interpreted the results and revised the manuscript. Dongmei Song: drew the figures. Linyuan Shen, Chunguang Zhao, Xiaopeng Yang and Yiying Yao: assisted data collection and checked data. Chenyang Xu, Chenchen Bi, Zhongkui Xion: assisted data collection.

---

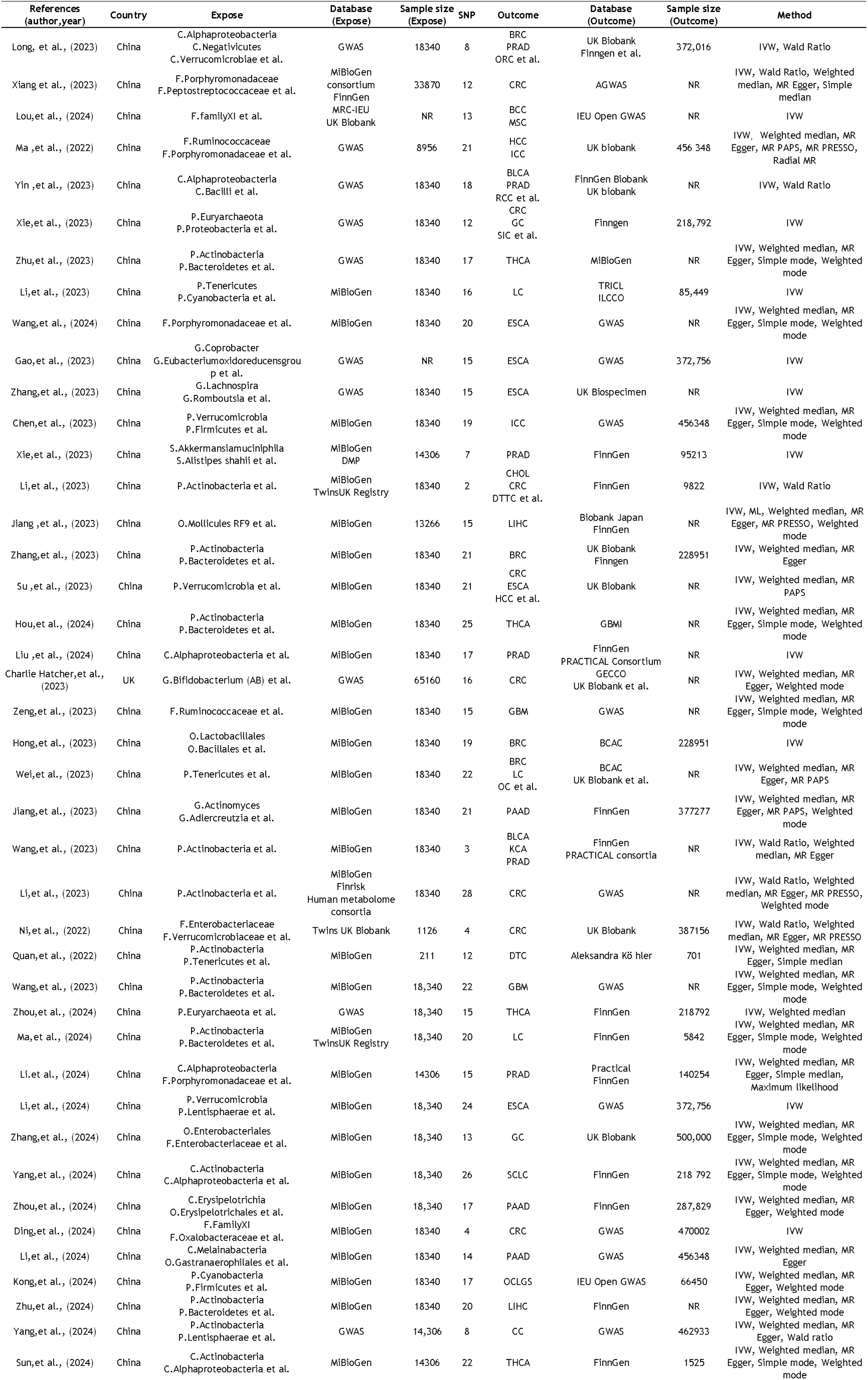
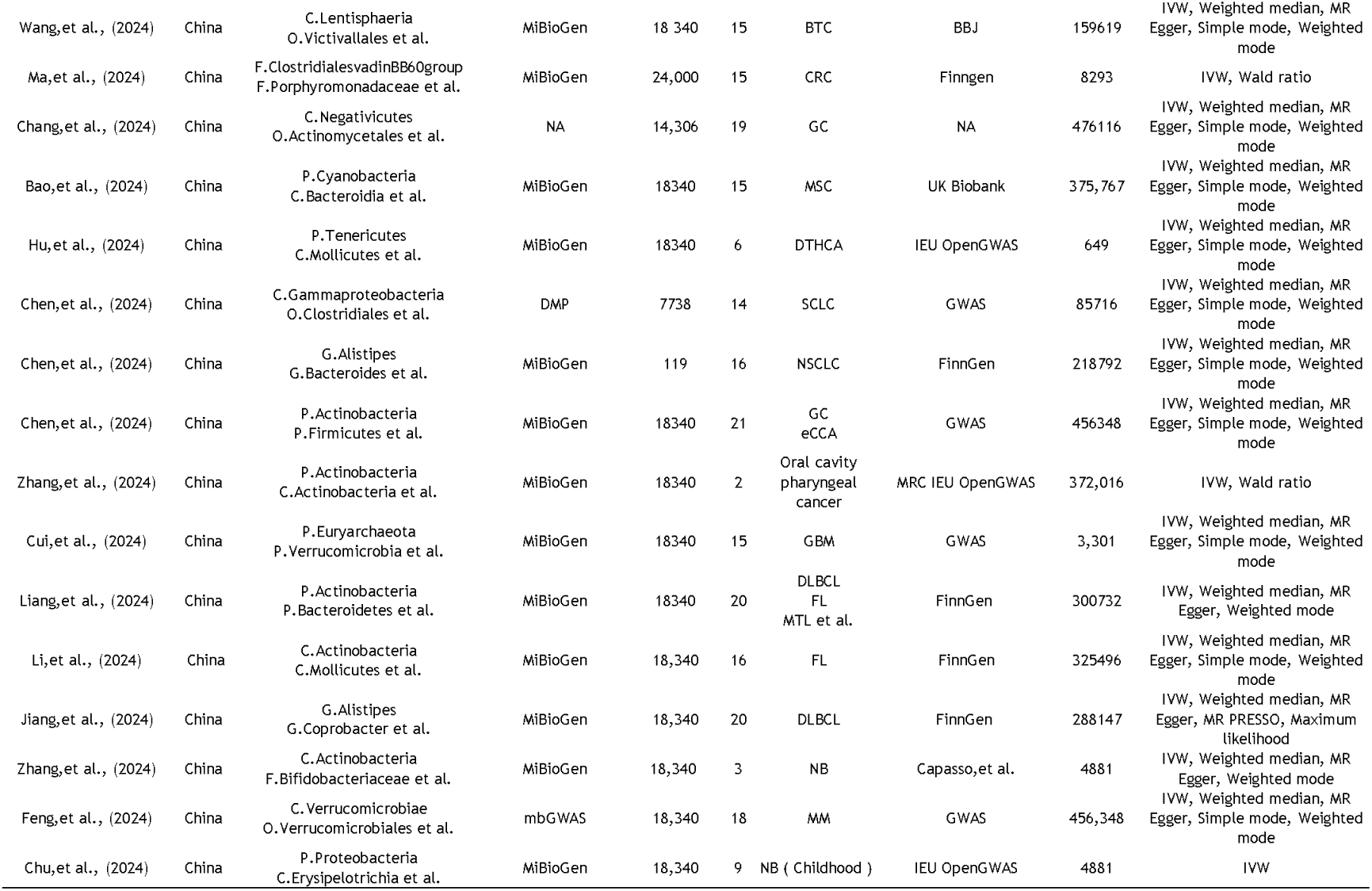

## References

1. Panebianco C, Andriulli A, Pazienza V. Pharmacomicrobiomics: exploiting the drug-microbiota interactions in anticancer therapies. MICROBIOME 2018; 6(1): 92.

2. Cao Y, Xia H, Tan X et al.. Intratumoural microbiota: a new frontier in cancer development and therapy. SIGNAL TRANSDUCT TAR 2024; 9(1): 15.

3. Hanahan D. Hallmarks of Cancer: New Dimensions. CANCER DISCOV 2022; 12(1): 31–46.

4. Ting NL, Lau HC, Yu J. Cancer pharmacomicrobiomics: targeting microbiota to optimise cancer therapy outcomes. GUT 2022; 71(7): 1412–1425.

5. Alexander JL, Wilson ID, Teare J et al.. Gut microbiota modulation of chemotherapy efficacy and toxicity. NAT REV GASTRO HEPAT 2017; 14(6): 356–365.

6. Shiao SL, Kershaw KM, Limon JJ et al.. Commensal bacteria and fungi differentially regulate tumor responses to radiation therapy. CANCER CELL 2021; 39(9): 1202–1213.

7. Stein-Thoeringer CK, Saini NY, Zamir E et al.. A non-antibiotic-disrupted gut microbiome is associated with clinical responses to CD19-CAR-T cell cancer immunotherapy. NAT MED 2023; 29(4): 906–916.

8. Sepich-Poore GD, Zitvogel L, Straussman R et al.. The microbiome and human cancer. SCIENCE 2021; 371(6536).

9. Delannoy-Bruno O, Desai C, Raman AS et al.. Evaluating microbiome-directed fibre snacks in gnotobiotic mice and humans. NATURE 2021; 595(7865): 91–95.

10. Fluhr L, Mor U, Kolodziejczyk AA et al.. Gut microbiota modulates weight gain in mice after discontinued smoke exposure. NATURE 2021; 600(7890): 713–719.

11. Wan Y, Wang F, Yuan J et al.. Effects of dietary fat on gut microbiota and faecal metabolites, and their relationship with cardiometabolic risk factors: a 6-month randomised controlled-feeding trial. GUT 2019; 68(8): 1417–1429.

12. Malard F, Vekhoff A, Lapusan S et al.. Gut microbiota diversity after autologous fecal microbiota transfer in acute myeloid leukemia patients. NAT COMMUN 2021; 12(1): 3084.

13. Larsson SC, Burgess S. Causal role of high body mass index in multiple chronic diseases: a systematic review and meta-analysis of Mendelian randomization studies. BMC MED 2021; 19(1): 320.

14. Davies NM, Holmes MV, Davey SG. Reading Mendelian randomisation studies: a guide, glossary, and checklist for clinicians. BMJ-BRIT MED J 2018; 362: k601.

15. Sanderson E, Glymour MM, Holmes MV et al.. Mendelian randomization. Nat Rev Methods Primers 2022; 2.

16. Smith GD, Ebrahim S. ’Mendelian randomization’: can genetic epidemiology contribute to understanding environmental determinants of disease? INT J EPIDEMIOL 2003; 32(1): 1–22.

17. Long Y, Tang L, Zhou Y et al.. Causal relationship between gut microbiota and cancers: a two-sample Mendelian randomisation study. BMC MED 2023; 21(1): 66.

18. Jiang P, Yu F, Zhou X et al.. Dissecting causal links between gut microbiota, inflammatory cytokines, and DLBCL: a Mendelian randomization study. BLOOD ADV 2024; 8(9): 2268–2278.

19. Xiang Y, Zhang C, Wang J et al.. Identification of host gene-microbiome associations in colorectal cancer patients using mendelian randomization. J TRANSL MED 2023; 21(1): 535.

20. Lou J, Cui S, Li J et al.. Causal relationship between the gut microbiome and basal cell carcinoma, melanoma skin cancer, ease of skin tanning: evidence from three two-sample mendelian randomisation studies. FRONT IMMUNOL 2024; 15: 1279680.

21. Li B, Han Y, Fu Z et al.. The causal relationship between gut microbiota and lymphoma: a two-sample Mendelian randomization study. FRONT IMMUNOL 2024; 15: 1397485.

22. Ma J, Li J, Jin C et al.. Association of gut microbiome and primary liver cancer: A two-sample Mendelian randomization and case–control study. LIVER INT 2023; 43(1): 221–233.

23. Xie N, Wang Z, Shu Q et al.. Association between Gut Microbiota and Digestive System Cancers: A Bidirectional Two-Sample Mendelian Randomization Study. NUTRIENTS 2023; 15(13).

24. Yin Z, Liu B, Feng S et al.. A Large Genetic Causal Analysis of the Gut Microbiota and Urological Cancers: A Bidirectional Mendelian Randomization Study. NUTRIENTS 2023; 15(18).

25. Li Y, Wang K, Zhang Y et al.. Revealing a causal relationship between gut microbiota and lung cancer: a Mendelian randomization study. FRONT CELL INFECT MI 2023; 13: 1200299.

26. Zhu F, Zhang P, Liu Y et al.. Mendelian randomization suggests a causal relationship between gut dysbiosis and thyroid cancer. FRONT CELL INFECT MI 2023; 13: 1298443.

27. Wang K, Wang S, Qin X et al.. The causal relationship between gut microbiota and biliary tract cancer: comprehensive bidirectional Mendelian randomization analysis. FRONT CELL INFECT MI 2024; 14: 1308742.

28. Zhang Q, Wang H, Tian Y et al.. Mendelian randomization analysis to investigate the gut microbiome in oral and oropharyngeal cancer. FRONT CELL INFECT MI 2023; 13: 1210807.

29. Liang J, Liu G, Wang W et al.. Causal relationships between gut microbiota and lymphoma: a bidirectional Mendelian randomization study. FRONT CELL INFECT MI 2024; 14: 1374775.

30. Li X, Liang Z. Causal effect of gut microbiota on pancreatic cancer: A Mendelian randomization and colocalization study. J CELL MOL MED 2024; 28(8): e18255.

31. Riquelme E, Zhang Y, Zhang L et al.. Tumor Microbiome Diversity and Composition Influence Pancreatic Cancer Outcomes. CELL 2019; 178(4): 795–806.

32. Laniewski P, Ilhan ZE, Herbst-Kralovetz MM. The microbiome and gynaecological cancer development, prevention and therapy. NAT REV UROL 2020; 17(4): 232–250.

33. Pushalkar S, Hundeyin M, Daley D et al.. The Pancreatic Cancer Microbiome Promotes Oncogenesis by Induction of Innate and Adaptive Immune Suppression. CANCER DISCOV 2018; 8(4): 403–416.

34. Knippel RJ, Drewes JL, Sears CL. The Cancer Microbiome: Recent Highlights and Knowledge Gaps. CANCER DISCOV 2021; 11(10): 2378–2395.

35. Derosa L, Routy B, Desilets A et al.. Microbiota-Centered Interventions: The Next Breakthrough in Immuno-Oncology? CANCER DISCOV 2021; 11(10): 2396–2412.

36. Roberti MP, Yonekura S, Duong C et al.. Chemotherapy-induced ileal crypt apoptosis and the ileal microbiome shape immunosurveillance and prognosis of proximal colon cancer. NAT MED 2020; 26(6): 919–931.

37. Goubet AG, Wheeler R, Fluckiger A et al.. Multifaceted modes of action of the anticancer probiotic Enterococcus hirae. CELL DEATH DIFFER 2021; 28(7): 2276–2295.

38. Griffin ME, Espinosa J, Becker JL et al.. Enterococcus peptidoglycan remodeling promotes checkpoint inhibitor cancer immunotherapy. SCIENCE 2021; 373(6558): 1040–1046.

39. Yoon HS, Cho CH, Yun MS et al.. Akkermansia muciniphila secretes a glucagon-like peptide-1-inducing protein that improves glucose homeostasis and ameliorates metabolic disease in mice. NAT MICROBIOL 2021; 6(5): 563–573.

40. Grajeda-Iglesias C, Durand S, Daillere R et al.. Oral administration of Akkermansia muciniphila elevates systemic antiaging and anticancer metabolites. Aging (Albany NY) 2021; 13(5): 6375–6405.

41. Fernandes MR, Aggarwal P, Costa RGF et al.. Targeting the gut microbiota for cancer therapy. Nature reviews. Cancer 2022; 22(12): 703–722.

42. Fluckiger A, Daillere R, Sassi M et al.. Cross-reactivity between tumor MHC class I-restricted antigens and an enterococcal bacteriophage. SCIENCE 2020; 369(6506): 936–942.

43. Dohlman AB, Arguijo MD, Ding S et al.. The cancer microbiome atlas: a pan-cancer comparative analysis to distinguish tissue-resident microbiota from contaminants. CELL HOST MICROBE 2021; 29(2): 281–298.

44. Wu AH, Tseng C, Vigen C et al.. Gut microbiome associations with breast cancer risk factors and tumor characteristics: a pilot study. BREAST CANCER RES TR 2020; 182(2): 451–463.

45. Sanapareddy N, Legge RM, Jovov B et al.. Increased rectal microbial richness is associated with the presence of colorectal adenomas in humans. ISME J 2012; 6(10): 1858–1868.

46. Routy B, Le Chatelier E, Derosa L et al.. Gut microbiome influences efficacy of PD-1-based immunotherapy against epithelial tumors. SCIENCE 2018; 359(6371): 91–97.

47. Mahlen SD, Clarridge JR. Site and clinical significance of Alloscardovia omnicolens and Bifidobacterium species isolated in the clinical laboratory. J CLIN MICROBIOL 2009; 47(10): 3289–3293.

48. Xiang K, Wang P, Xu Z et al.. Causal Effects of Gut Microbiome on Systemic Lupus Erythematosus: A Two-Sample Mendelian Randomization Study. FRONT IMMUNOL 2021; 12: 667097.

49. Crost EH, Coletto E, Bell A et al.. Ruminococcus gnavus: friend or foe for human health. FEMS MICROBIOL REV 2023; 47(2).

50. Sivan A, Corrales L, Hubert N et al.. Commensal Bifidobacterium promotes antitumor immunity and facilitates anti-PD-L1 efficacy. SCIENCE 2015; 350(6264): 1084–1089.

51. Tao J, Li S, Gan RY et al.. Targeting gut microbiota with dietary components on cancer: Effects and potential mechanisms of action. CRIT REV FOOD SCI 2020; 60(6): 1025–1037.

52. Yang J, Yu J. The association of diet, gut microbiota and colorectal cancer: what we eat may imply what we get. PROTEIN CELL 2018; 9(5): 474–487.

53. Cunningham M, Azcarate-Peril MA, Barnard A et al.. Shaping the Future of Probiotics and Prebiotics. TRENDS MICROBIOL 2021; 29(8): 667–685.

54. Inamura K, Hamada T, Bullman S et al.. Cancer as microenvironmental, systemic and environmental diseases: opportunity for transdisciplinary microbiomics science. GUT 2022.

55. Yang L, Li A, Wang Y et al.. Intratumoral microbiota: roles in cancer initiation, development and therapeutic efficacy. SIGNAL TRANSDUCT TAR 2023; 8(1): 35.

